# Diagnostic accuracy, operational feasibility, and cost considerations of Truenat MTB Plus and Xpert MTB/RIF Ultra for pulmonary tuberculosis

**DOI:** 10.64898/2026.01.19.26344389

**Authors:** Zeeshan Sidiq, Ankita Anand, Kaushal Kumar Dwivedi, Payal Tyagi, Sanjay Rajpal, Kamal Kishore Chopra, Shikha Dhawan, Vishal Khanna

**Affiliations:** New Delhi Tuberculosis Centre, JLN Marg, Delhi Gate, New Delhi, Delhi, India; Centre for Social Integration and borderless world, Ghaziabad, Uttar Pradesh, India; Chest Clinic Lok Nayak Hospital, Lok Nayak Jai Prakash Hospital, JLN Marg, Delhi Gate, New Delhi, Delhi, India

**Author notes:** These authors contributed equally to this work.

**Keywords:** Tuberculosis, Truenat MTB Plus, Xpert MTB/RIF Ultra, Diagnostic accuracy, Decentralized diagnostics

## Abstract

**Background:** Tuberculosis (TB) remains a major global health challenge, necessitating rapid and accurate diagnostic tools. The World Health Organization (WHO) endorses nucleic acid amplification tests (NAATs) like Truenat MTB Plus and Xpert MTB/RIF Ultra. While both are widely implemented, direct comparisons integrating diagnostic accuracy with operational and cost considerations across real-world settings are limited. This study aimed to compare the diagnostic accuracy of Truenat MTB Plus and Xpert MTB/RIF Ultra for the detection of *Mycobacterium tuberculosis* (MTB) and rifampicin resistance (RIF-R) among presumptive pulmonary TB patients in India, while also assessing their operational feasibility and cost-related parameters.

**Methods:** This prospective diagnostic accuracy study was conducted at New Delhi Tuberculosis Centre, in New Delhi, India, enrolling 664 presumptive pulmonary TB patients. Paired sputum specimens were tested using Truenat MTB Plus/MTB-RIF and Xpert MTB/RIF Ultra. Mycobacterial culture was used as the primary reference standard for MTB detection, and phenotypic drug susceptibility testing (pDST) was used for RIF-R detection. Sensitivity, specificity, and overall accuracy were calculated, and operational parameters, including invalid/indeterminate rates and cost per test, were recorded.

**Results:** The overall prevalence of culture-confirmed TB was 14.8% (98/664), with a RIF-R prevalence of 7.1% (7/98). Compared with culture, Xpert MTB/RIF Ultra demonstrated a sensitivity of 97.9% (95% CI: 95.2–100%) and a specificity of 95.5% (95% CI: 93.7–97.3%). Truenat MTB Plus showed a sensitivity of 92.5% (95% CI: 85.1–96.6%) and a specificity of 90.1% (95% CI: 86.8–92.6%). For RIF-R detection, Xpert Ultra had a sensitivity of 83.3% and a specificity of 100%, compared to Truenat MTB-RIF’s sensitivity of 75.0% and specificity of 100%.

Operationally, Truenat had a significantly higher rate of invalid results (5.9% vs 0%) and RIF indeterminate results (46.5% vs 13.2%) compared to Xpert Ultra. However, the cost per test for Truenat was substantially lower (approx. INR 800 for MTB Plus vs INR 1800 for Ultra). Truenat’s minimal infrastructure requirements make it highly suitable for peripheral and district-level decentralized testing, while Xpert Ultra is better suited for district and reference laboratories.

**Conclusion:** Both Truenat MTB Plus and Xpert MTB/RIF Ultra demonstrated excellent diagnostic accuracy for TB detection, significantly outperforming smear microscopy. Xpert Ultra showed superior accuracy and lower indeterminate rates, but Truenat offers substantial operational and cost advantages for decentralized deployment. These findings support a tiered diagnostic approach, leveraging the strengths of both platforms to expand molecular testing access and accelerate progress toward global TB elimination goals.

## Introduction

Tuberculosis (TB) remains a major global public health challenge, requiring timely and accurate diagnostic tools to enable early treatment and reduce transmission. In 2023, an estimated 10.8 million people developed TB globally, yet only 8.2 million cases were notified, leaving approximately 2.7 million individuals undiagnosed or unreported. Among those diagnosed, fewer than half (48%) received upfront testing with a World Health Organization (WHO)– recommended rapid molecular assay, while the remainder were evaluated using less sensitive methods such as smear microscopy (1).

Conventional TB diagnostics have important limitations. Smear microscopy has low sensitivity, particularly in paucibacillary disease, whereas culture, although highly specific, requires prolonged turnaround times. To address these gaps, WHO recommends nucleic acid amplification tests (NAATs) as initial diagnostic tools, as they provide improved sensitivity, rapid results, and the ability to detect drug resistance–associated mutations.

Truenat MTB/MTB Plus (Molbio Diagnostics) and Xpert MTB/RIF Ultra (Cepheid) are among the most widely implemented WHO-endorsed NAAT platforms. Truenat is a chip-based, real-time PCR system designed for decentralized testing. The Truenat MTB Plus assay incorporates dual targets (IS6110 and IS1081) to enhance detection of *Mycobacterium tuberculosis*, particularly in regions where strains with low IS6110 copy numbers are prevalent. In contrast, Xpert MTB/RIF Ultra is a fully automated cartridge-based assay with improved sensitivity in smear-negative and paucibacillary TB, although concerns regarding reduced specificity and infrastructure requirements persist.

Although several studies have evaluated these assays individually, direct head-to-head comparisons that integrate diagnostic accuracy with operational feasibility and cost-related considerations across different healthcare settings remain limited.

Therefore, this study aimed to compare the diagnostic accuracy of Truenat MTB Plus and Xpert MTB/RIF Ultra among presumptive pulmonary TB patients using mycobacterial culture as the reference standard, while also assessing their operational feasibility, cost-effectiveness, and scalability.

## Methods

### Study design

This prospective diagnostic accuracy study assessed the performance of Xpert MTB/RIF Ultra and Truenat MTB Plus/MTB-RIF at the New Delhi Tuberculosis Centre in New Delhi, India. Participant recruitment for this study was conducted from 04/07/2024 to 27/12/2024. The study population consisted of patients who presented to the outpatient department of the Chest Clinic, Lok Nayak Jaiprakash Hospital, with a clinical suspicion of pulmonary tuberculosis. Participants were consecutively enrolled if they fulfilled the following criteria: (i) provided written informed consent, (ii) had no prior history of anti-tuberculosis therapy, and (iii) were able to submit adequate quality sputum specimens.

The study aimed to recruit approximately 1000 participants to obtain at least 100 Mycobacterium tuberculosis (MTB)–positive specimens and a minimum of 20 rifampicin-resistant (RIF-R) specimens. The sample size was calculated with α = 0.05 and 80% power based on reported sensitivities of 0.88 for Xpert MTB/RIF Ultra and 0.73 for Truenat MTB Dx (2). Enrollment was concluded once the predefined recruitment targets had been achieved. This study adhered to the ethical principles outlined in the Declaration of Helsinki (1964) and its subsequent revisions. Approval was obtained from the appropriate institutional scientific and ethics committees.

### Procedures

The enrolled participants were requested to provide two consecutive sputum specimens, which were promptly transported to the laboratory. The paired samples were pooled, homogenized, and divided into three equal aliquots. One aliquot was tested using the Xpert MTB/RIF Ultra assay, the second with the Truenat MTB Plus assay, and the third was used for smear microscopy and mycobacterial culture. Pooling was performed to ensure that all diagnostic tests were conducted on identical material, thereby enabling a direct and reliable comparison of assay performance. All procedures for smear microscopy, liquid culture, Drug Susceptibility testing (DST), Truenat, and GeneXpert were performed following standard protocols (3,4,5,6,7). Liquid culture was used as the primary reference standard for evaluating the diagnostic accuracy of Truenat MTB Plus and Xpert MTB/RIF Ultra, in line with WHO recommendations. Sensitivity, specificity, positive predictive value (PPV), negative predictive value (NPV), and overall accuracy were calculated using culture-positive and culture-negative specimens. A composite reference standard (CRS)—defined as positivity by either liquid culture or smear microscopy—was applied as a secondary analysis to account for potential false-negative culture results; CRS-based estimates were considered supportive. Phenotypic drug susceptibility testing (pDST) on culture-positive isolates served as the reference standard for rifampicin resistance detection. Indeterminate rifampicin resistance results were excluded from accuracy calculations and analyzed separately for operational implications. 95% confidence intervals were calculated using the Wilson method, and rifampicin resistance analyses were considered exploratory due to the limited number of resistant cases.

*Note: Diagnostic accuracy analyses were performed using only samples for which valid results were available for both the index test and the corresponding reference standard. Consequently, denominators varied across comparisons due to invalid molecular test results, contaminated cultures, or unavailability of phenotypic drug susceptibility testing*.

## Results

During the study period, a total of 664 participants were included in the study, comprising 382 males (57.5%) and 282 females (42.5%), with a male-to-female ratio of approximately 1.35:1. The participants’ ages ranged from 10 to 91 years, with a median age of 36.5 years. The largest representation was from the 36–59 years group (240/664, 36.1%), followed closely by the 19– 35 years group (217/664, 32.7%). Overall, the cohort was predominantly adult, with substantial representation of both pediatric and elderly populations, allowing for a robust demographic analysis. A total of 98 patients were culture-confirmed for *Mycobacterium tuberculosis*, giving an overall Tuberculosis prevalence of 14.8%. Rifampicin resistance was detected in seven cases, corresponding to an overall prevalence of 7.1%. The detailed results are given in Table 1

**Table 1:** Age- and Sex-wise Distribution of TB and Rifampicin Resistance.

| Age Group | Male (n) | Female (n) | Total (n) | % of total | Culture-positive TB (n) | TB prevalence (%) | Rifampicin-resistant TB (n) | RR-TB prevalence (%) |
| --- | --- | --- | --- | --- | --- | --- | --- | --- |
| ≤18 | 51 | 51 | 102 | 15.4 | 17 | 16.7 | 1 | 5.9 |
| 19–35 | 107 | 110 | 217 | 32.7 | 35 | 16.1 | 2 | 5.7 |
| 36–59 | 145 | 95 | 240 | 36.1 | 35 | 14.6 | 3 | 8.6 |
| ≥60 | 79 | 26 | 105 | 15.8 | 11 | 10.5 | 1 | 9.1 |
| **Total** | **382** | **282** | **664** | **100** | **98** | **14.7** | **7** | **7.1** |

### Diagnostic Accuracy of Truenat MTB Plus assay

Truenat MTB Plus demonstrated a sensitivity of 98.4% (95% CI: 91.7–99.9%) and specificity of 85.3% (95% CI: 82.0–88.2%) against microscopy (N = 625), with a PPV of 42.4%, NPV of 99.8%, and an overall accuracy of 86.0%. Compared with culture (N = 596), the sensitivity and specificity were 92.5% (85.1–96.6%) and 90.1% (86.8–92.6%), respectively, with PPV 63.2%, NPV 98.5%, and accuracy 90.4%. Against a composite reference standard (smear or culture; N = 625), MTB Plus achieved 92.8% sensitivity (85.6–96.6%), 89.8% specificity (86.4–92.4%), PPV 62.5%, NPV 98.5%, and an accuracy of 90.2%. For rifampicin resistance detection, MTB-RIF compared with phenotypic DST (N = 64) showed 75.0% sensitivity (30.1–95.4%), 100% specificity (93.9– 100%), PPV 100%, NPV 98.4%, and an overall accuracy of 98.4%. The results are presented in Table 2 and figure 1.

**Table 2:**
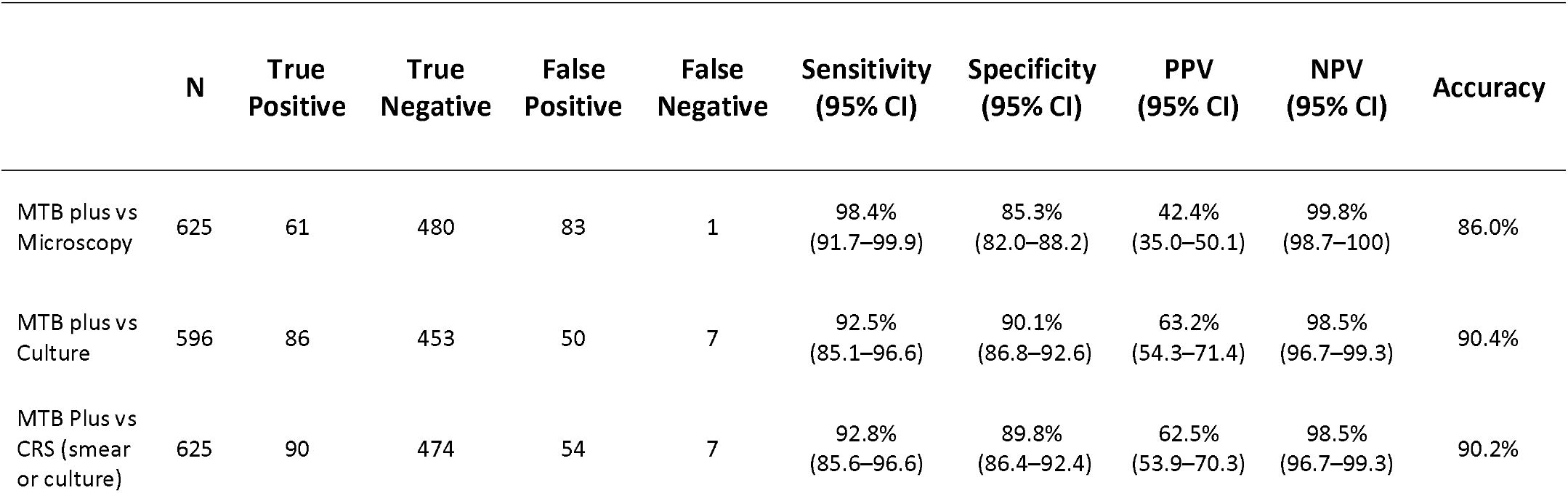
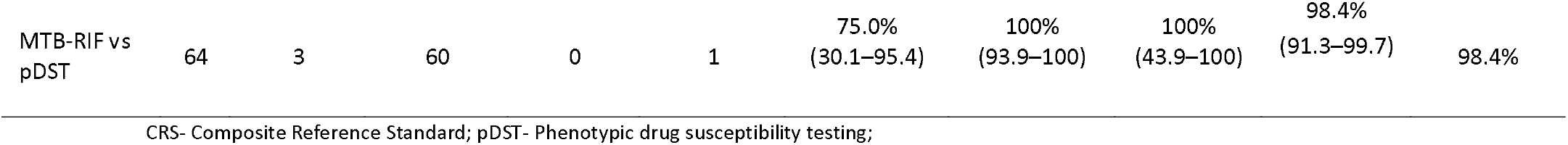
Diagnostic performance of Truenat MTB Plus and Truenat MTB-RIF Dx compared with microscopy, culture, and composite reference standard and pDST for tuberculosis and rifampicin resistance detection.

|  | N | True Positive | True Negative | False Positive | False Negative | Sensitivity (95% CI) | Specificity (95% CI) | PPV (95% CI) | NPV (95% CI) | Accuracy |
| --- | --- | --- | --- | --- | --- | --- | --- | --- | --- | --- |
| MTB plus vs Microscopy | 625 | 61 | 480 | 83 | 1 | 98.4% (91.7–99.9) | 85.3% (82.0–88.2) | 42.4% (35.0–50.1) | 99.8% (98.7–100) | 86.0% |
| MTB plus vs Culture | 596 | 86 | 453 | 50 | 7 | 92.5% (85.1–96.6) | 90.1% (86.8–92.6) | 63.2% (54.3–71.4) | 98.5% (96.7–99.3) | 90.4% |
| MTB Plus vs CRS (smear or culture) | 625 | 90 | 474 | 54 | 7 | 92.8% (85.6–96.6) | 89.8% (86.4–92.4) | 62.5% (53.9–70.3) | 98.5% (96.7–99.3) | 90.2% |
| MTB-RIF vs<br>pDST | 64 | 3 | 60 | 0 | 1 | 75.0%<br>(30.1–95.4) | 100%<br>(93.9–100) | 100%<br>(43.9–100) | 98.4%<br>(91.3–99.7) | 98.4% |

CRS- Composite Reference Standard; pDST- Phenotypic drug susceptibility testing;

### Diagnostic Accuracy of the Xpert MTB/RIF Ultra Assay

Against microscopy (N=663), Xpert MTB/RIF Ultra showed a sensitivity of 98.5% (95% CI: 91.9– 99.8%) and specificity of 89.4% (95% CI: 86.7–91.8%), with a PPV of 51.2% and NPV of 99.8%, yielding an accuracy of 90.3 %. Compared with culture (N = 633), the sensitivity and specificity were 97.9% (95% CI: 95.2–100%) and 95.5% (95% CI: 93.7–97.3%), respectively, with an accuracy of 95.9%. Against the composite reference standard (smear or culture, N = 663), the sensitivity was 98.0% (95% CI: 93.1–99.8%) and specificity was 94.8% (95% CI: 92.7–96.5%), achieving 95.3% accuracy. Xpert MTB/RIF Ultra versus phenotypic DST (N = 93) for rifampicin resistance detected showed 83.3% sensitivity (95% CI: 43.6–97.0%) and 100% specificity (95% CI: 95.8–100%), with an accuracy of 98.9%. The results are detailed in Table 3.

**Table 3:** Diagnostic performance of Xpert MTB/RIF Ultra compared with microscopy, culture, and composite reference standard and pDST for tuberculosis and rifampicin resistance detection.

|  | N | True<br>Positive | True<br>Negative | False<br>Positive | False<br>Negative | Sensitivity<br>(95% CI) | Specificity<br>(95% CI) | PPV (95%<br>CI) | NPV (95% CI) | Accuracy |
| --- | --- | --- | --- | --- | --- | --- | --- | --- | --- | --- |
| Xpert vs<br>Microscopy | 663 | 66 | 533 | 63 | 1 | 98.5%<br>(91.9–99.8) | 89.4%<br>(86.7–91.8) | 51.2.4%<br>(42.2–60.1) | 99.8%<br>(99.0–100) | 90.3% |
| Xpert vs<br>Culture | 633 | 96 | 511 | 24 | 2 | 97.9%<br>(95.2–100) | 95.5%<br>(93.7–97.3) | 80.0%<br>(72.8–87.2) | 99.6%<br>(99.1–100) | 95.9% |
| Xpert vs<br>CRS (smear<br>or culture) | 663 | 100 | 532 | 29 | 2 | 98.0%<br>(93.1–99.8) | 94.8%<br>(92.7–96.5) | 77.5%<br>(69.3–84.4) | 99.6%<br>(98.7–99.9) | 95.3% |
| RIF Ultra vs<br>pDST | 93 | 5 | 87 | 0 | 1 | 83.3%<br>(43.6–97.0) | 100%<br>(95.8–100) | 100%<br>(56.6–100) | 98.9%<br>(93.8–99.8) | 98.9% |

CRS- Composite Reference Standard; pDST- Phenotypic drug susceptibility testing

**Table 3.** Comparison of Operational and Cost-related Parameters Between Truenat MTB Plus / RIF and Xpert MTB/RIF Ultra.

| Parameter | Truenat MTB Plus / RIF | Xpert MTB/RIF Ultra |
| --- | --- | --- |
| Cost per test (approx., INR) | 800 + 562* | 1800 |
| Instrument cost | Low–Moderate | High |
| MTB invalid results | 39/625 (6.2%) | 0/625 (0%) |
| RIF indeterminate results | 67/144 (46.5%) | 17/129 (13.2%) |
| Cartridge clogging | 74 instances | 0 |
| Instrument errors | 13 | 2 (5006, 5011) |
| Repeat testing required | 52/625 (8.3%) | 2/625 (0.3%) |
| Average turnaround time | 60–90 min/sample | ~90 min/sample |
| Throughput (samples/run) | 1–4 | 1–4 |
| Infrastructure needs | Minimal | Moderate–High |
| Suitable level of use | Peripheral / District | District / Reference |

\*800 for MTB Plus and additional 562 for RIF chip

## Errors, Invalids and indeterminate results

### Sample Processing and Extraction-related Issues

During sample processing, occasional challenges were noted. Some samples failed to liquefy within the stipulated time despite extended or overnight incubation, while 74 instances of cartridge clogging were observed during DNA extraction with the Trueprep system. Though infrequent, these events affected workflow efficiency and sample quality. In contrast, with the Xpert MTB/RIF Ultra assay, only two errors (codes 5006 and 5011) were observed.

### Detection Errors in MTB Assays

A total of 39 (5.9%) invalid results were obtained for *Mycobacterium tuberculosis* detection using the Truenat MTB Plus assay, whereas no invalid results were observed with Xpert MTB/RIF Ultra. A single repeat testing was performed for all invalid samples, after which 3 (7.7%) yielded valid results while the remaining 36 (92.3%) persisted as invalid.

### Indeterminate Results for Rifampicin Resistance Detection

Among the samples that tested positive for *Mycobacterium tuberculosis*, 46.5% (67 out of 144) from the Truenat MTB Plus assay yielded indeterminate results for rifampicin resistance when tested with Truenat MTB-RIF. In comparison, 17 out of 129 (13.2%) MTB-positive samples tested by Xpert MTB/RIF Ultra were indeterminate for rifampicin resistance. Notably, all 17 indeterminate results on Xpert MTB/RIF Ultra corresponded to trace-positive MTB detections, whereas the Truenat MTB Plus assay showed indeterminate outcomes across a range of MTB detection grades, from very low to high.

## Operational Feasibility and Cost-related Parameters

Operational and cost-related parameters were evaluated to assess assay feasibility across different healthcare settings. Key factors included cost per test, infrastructure requirements, turnaround time, and the frequency of invalid or indeterminate results. Data on assay throughput, infrastructure needs, and cost per test were recorded to support the subsequent analysis of cost-effectiveness and scalability. The results are detailed in table 3.

## Discussion

This study offers a detailed, real-world comparison of two WHO-endorsed molecular diagnostic tools, Truenat MTB Plus and Xpert MTB/RIF Ultra, for the detection of *Mycobacterium tuberculosis* and rifampicin resistance among individuals with presumptive pulmonary TB in India. Both platforms performed well overall; however, important distinctions in accuracy, operational reliability, and error rates were observed that are relevant to national TB control efforts and the global drive for universal access to rapid drug-resistant testing.

Compared with culture as the reference standard, Xpert MTB/RIF Ultra achieved slightly higher sensitivity and specificity (97.9% and 95.5%, respectively) than Truenat MTB Plus (92.5% and 90.1%, respectively). This modest but consistent advantage mirrors the results of earlier multicentric evaluations that reported similar findings in Indian and African settings (8, 9,10). The improved analytical sensitivity of Ultra likely stems from its larger reaction volume, refined probe chemistry, and algorithm optimized for low bacillary load samples.

Both assays clearly outperformed smear microscopy, confirming the superiority of molecular testing for TB diagnosis of TB. Their high negative predictive values (≥98%) indicate that either test can reliably exclude disease in symptomatic individuals, which is an important consideration for screening and triage in high-burden programs.

Reliable detection of rifampicin resistance is essential to ensure that patients receive effective therapy early. In this evaluation, both assays demonstrated perfect specificity compared with phenotypic drug susceptibility testing, confirming that false-positive resistance calls were rare. However, Ultra showed higher sensitivity (83.3%) than Truenat MTB-RIF (75.0%) and a markedly lower proportion of indeterminate results (13.2% vs. 46.5%). Indeterminate rifampicin resistance results necessitate repeat testing, referral to higher-tier laboratories, or delayed treatment decisions, all of which can impact patient management and laboratory workload. All ultra-indeterminate results occurred in samples with trace-positive MTB detections, consistent with the algorithmic safeguards to prevent spurious resistance reporting in very low bacterial loads (5). In contrast, Truenat produced indeterminate outcomes across all detection categories, suggesting variability related to amplification or internal control thresholds. Comparable findings have been described in previous Truenat evaluations (7,8,11,12), where the multi-step extraction plus amplification workflow may have contributed to reduced assay stability. In contrast, Ultra’s integrated closed-cartridge design limits operator handling and potential DNA loss, which likely explains its greater consistency across sample types (7, 13, 14).

From a programmatic standpoint, both technologies offer clear advantages but serve different operational needs. Truenat, with its small footprint and portability, is well-suited for decentralized or peripheral deployment, allowing same-day results and faster treatment initiation in hard-to-reach areas. Ultra, which offers superior analytical precision and minimal manual intervention, is best positioned for district or reference laboratories where confirmatory testing, surveillance, and quality monitoring are conducted. This complementary approach aligns with the WHO’s current guidance that molecular platforms should be deployed according to local infrastructure and case load to maximize access and maintain high diagnostic standards (6). Therefore, a network that integrates both systems could ensure wider diagnostic coverage while preserving analytical rigor.

The main strengths of this investigation are its prospective design, use of identical pooled sputum for both assays, eliminating inter-sample variability, and adherence to standardized WHO and Global Laboratory Initiative procedures. These factors increase the comparability with other datasets and enhance confidence in the findings.

This study has certain limitations. The number of rifampicin-resistant cases identified during the study period was limited, and therefore estimates related to rifampicin resistance detection should be interpreted with caution. The study was conducted at a single high-volume diagnostic centre, which may limit generalizability to settings with lower testing throughput. Indeterminate results were excluded from diagnostic accuracy calculations; however, these were analyzed separately to highlight their operational implications. Finally, the cost analysis was descriptive in nature and intended to provide comparative insights rather than a formal economic evaluation.

In conclusion, both Truenat MTB Plus and Xpert MTB/RIF Ultra demonstrated excellent diagnostic accuracy for detecting Mycobacterium tuberculosis and rifampicin resistance. Xpert MTB/RIF Ultra showed higher sensitivity, specificity, and reproducibility, particularly for rifampicin resistance detection, with significantly fewer indeterminate results. Truenat MTB Plus, while slightly less precise, offers clear operational advantages through its portability, lower cost, and minimal infrastructure requirements, making it highly suitable for decentralized testing. Together, these findings support a tiered diagnostic approach—deploying Truenat to expand molecular testing at peripheral and district levels and Ultra at higher-tier laboratories for confirmatory testing—to strengthen diagnostic networks, improve access to rapid drug-resistance testing, and advance progress toward global TB elimination goals.

## Data Availability

All data produced in the present study are available upon reasonable request to the authors

## Notes

### Competing Interest Statement

The authors have declared no competing interest.

### Funding Statement

This study did not receive any funding

### Author Declarations

The Institutional Ethics Committee (IEC) of New Delhi Tuberculosis Centre has reviewed and approved the research proposal titled: "Comparison of New Molecular Diagnostics Xpert MTB/RIF Ultra and Truenat MTB Plus with Truenat MTB and Truenat MTB Rif for the detection of Mycobacterium tuberculosis and Rifampicin resistance".

